# A Genomic Snapshot of *Enterococcus faecalis* within Public Hospital Environments in South Africa

**DOI:** 10.1101/2022.03.07.22272060

**Authors:** Christiana O. Shobo, Daniel G. Amoako, Mushal Allam, Arshad Ismail, Sabiha Y. Essack, Linda A. Bester

## Abstract

Enterococci are among the most common opportunistic hospital pathogens. This study used whole-genome sequencing (WGS) and bioinformatics to determine the antibiotic resistome, genetic support, clones and phylogenetic relationship of *Enterococcus faecalis* isolated from hospital environments in South Africa. Isolates were recovered from 11 frequently touched sites by patients and healthcare workers in different wards at 4 levels of healthcare (A, B, C and D) in Durban, South Africa. Following microbial identification and antibiotic susceptibility tests. Of the 245 *E. faecalis* isolates identified, 38 were subjected to WGS on the Illumina MiSeq platform. The *tet(M)* (31/38, 82%) and *erm(C)* (16/38, 42%) genes were the most common antibiotic resistome found in isolates originating from the different hospital environments which corroborated with their antibiotic resistance phenotypes. The isolates harboured mobile genetic elements consisting of plasmids (n=11) and prophages (n=14), that were mostly clone-specific. Of note, a large number insertion sequence (IS) families were found with the IS3 (55%), IS5 (42%), IS1595 (40%) and Tn*3* Transposon been the most predominate. Microbial typing using WGS data revealed 15 clones with 6 major sequence types (ST) belonging to ST16 (n =7), ST40 (n = 6), ST21 (n =5), ST126 (n = 3), ST23 (n =3) and ST386 (n=3). Phylogenomic analysis showed that the major clones were mostly conserved within specific hospital environments. However, further metadata insights revealed the complex intra-clonal spread of these *E. faecalis* major clones between the sampling sites within each specific hospital setting. The results of these genomic analyses will offer insights into *E. faecalis* in the hospital environments relevant in the design of optimal infection prevention strategies in hospital settings.

## 1.0. Introduction

The surveillance of hospital environments can be a useful tool to better understand the opportunistic microbial communities within the hospital [1], to identify the source of an outbreak[2], and to evaluate the efficacy of environmental disinfection or other infection prevention and control measures [3]. Inadequate control practices have played a significant role in the dissemination, persistence, intra- and inter-hospital spread of drug-resistant organisms. Regrettably, good clinical trials comparing the different approaches to, and the impact of infection prevention and control interventions on the control of drug-resistant bacteria in hospitals and other healthcare facilities are minimal [4,5]. Accurate identification of resistant bacterial reservoirs and modes of transmission help inform such interventions.

The latest successes in tracing worldwide epidemics [6] and nosocomial outbreaks [7] have been attributed to whole-genome sequencing (WGS). Genomic comparison has aided our understanding of the evolution and spread of infectious agents. Comparative genomic analyses have been made possible through the use of WGS, showing the extent of genomic variation, which may result in varied phenotypes, thus expanding our understanding of diverse genomic determinants such as antibiotic resistance genes and their genetic support in bacterial species [7,8].

*Enterococcus faecalis (E. faecalis)* is a good indicator bacterium in hospital environment monitoring, being Gram-positive cocci these opportunistic pathogens not only form noxious biofilms on implanted medical devices and catheters, they also cause abdominal infection, urinary tract infections, surgical site wound infections, bacteremia, endocarditis and burn [9]. Antibiotic resistance is either intrinsic or through sporadic mutation or through the acquisition of foreign genetic material, by horizontal gene exchange occurring with the aid of mobile genetic elements plasmids, prophages and insertion sequences [10,11]. Difficulties in treating *E. faecalis* and *E. faecium* (most prevalent species in human) have emerged due to acquired resistance, predominantly multi-drug resistance to universally used drugs as well as vancomycin [12]. A number of previous surveillance studies involving *E. faecalis* in Africa have focused either on wastewater treatment plants (WWTPs) and hospital effluent but not on the internal hospital environment [13,14]. Moreover, in South Africa, studies on the contamination of *E. faecalis*, using high discrimination resolution typing are scarce. This study, therefore, uses WGS in delineating the resistome, mobile genetic support, the clones and phylogenomic relationship of *E. faecalis* isolated from the hospital environment in places frequently touched by patients and healthcare workers at four different levels of healthcare in the metropolitan city of Durban, South Africa.

## 2.0. Materials and Methods

### 2.1. Ethical approval

Ethical clearance was received from the Biomedical Research Ethics Committee of the University of KwaZulu-Natal (Ref. BE606/16). The study was also registered on the Health Research and Knowledge Management database (HRKM 098/17) of the KwaZulu-Natal Provincial Health Research Ethics Committee. Gate keeper ‘s approval was further granted by participating hospitals. No Human samples were taken in the study. All protocols were executed according to the agreed ethical approval terms and conditions.

### 2.2. Study setting

The selected hospitals were all public hospitals situated in the eThekwini region in Durban, South Africa. For non-disclosure reasons, the names of the hospitals were withheld and referred to as A, B, C and D representing central, tertiary, regional and district facilities, respectively. The central hospital (A), with a 1200 bed-size, offers tertiary level sub-specialist services and serves as a referral hospital for the district, regional and tertiary hospitals. The tertiary hospital (B) with 800 beds also has specialist services and receives referrals from regional and district hospitals not limited to provincial boundaries. The regional hospital (C), with a 743 bed-size, provides services to a specific regional population and receives referrals from several district hospitals. The district hospital (D) has 300 beds and serves as a health district and supports primary health care services on a 24-hour basis. Samples were collected in the intensive care unit (ICU) and paediatric ward from 11 sites that included the telephones, ventilators, blood pressure apparatus, patient files, drip stands, sinks, occupied beds, unoccupied beds, nurses’ tables, mops and the door handle of the linen room. A total number of 620 samples were collected over a period of three months from the four levels of healthcare. These samples were collected weekly in batches viz; 1^st^ batch collected at the beginning of the week, 2^nd^ in the middle of the week and 3^rd^ batch at the end of the week. All samples were collected by randomly swabbing approximately 5 cm of the site using pre-labelled Nylon flock swabs with transport media (FLOQSwabs COPEN diagnostics Inc, USA). The swabs were then transported to the laboratory in iceboxes and processed within 3 to 4 hrs of sampling.

### 2.3. Isolation and identification of *Enterococcus*

#### 2.3.1. Phenotypic determination of *Enterococcus*

The samples were inoculated separately into Tryptone Soya Broth (TSB) (Oxoid, Hampshire, England) and incubated at 37 °C for 2 h with shaking at 100 rpm. Following incubation, 1 ml of each culture was inoculated into 9 ml of TSB supplemented with 6.5% NaCl and incubated at 37 °C for 24 h with shaking at 100 rpm. All 24 h cultures were sub-cultured by spread plating 100 µl onto Bile Esculin Azide agar (Himedia, Mumbai, India). Plates were incubated for 24 h at 37 °C, and brown-grey colonies surrounded by black halos were considered presumptive enterococci. Presumptive colonies were streaked onto Bile Aesculin agar (Lab M, Lancashire, UK), and incubated at 37 °C to obtain pure colonies. For characterisation of haemolysis, cultures were prepared on 5% Sheep Blood agar (Oxoid, Hampshire, England), and on Tryptone Soya Agar (TSA) (Oxoid, Hampshire, England) for biochemical characterisation and the Gram stain test [15]. Phenotypic identification was undertaken using API 20 Strep kits (Biomerieux SA, Marcy I ‘Etoile, France). *Staphylococcus aureus* American Type Culture Collection (ATCC) 29213 and *E. faecalis* ATCC 29212 were used as controls. Presumptive enterococci were stored in 10% glycerol stock solution at - 80 °C until further processing.

#### 2.3.2. Molecular confirmation of isolates

Stock cultures were resuscitated on TSA plates incubated at 37 °C for 24 h. DNA was extracted using the heat lysis method as previously described [16]. A multiplex polymerase chain reaction (PCR) was performed to confirm isolates at the genus and species level. Genus-specific and species-specific primer used in all the reactions were as previously described [17,18] **(Table S1)**. Two PCR reaction mixtures, both containing the *Enterococcus* genus-specific primers, were set up for different primer sets as follows: Group 1: *E. faecalis*. Each reaction was performed in a total volume of 15 µl consisting of 8 µl of DreamTaq Green PCR Master Mix (2X) (Thermo Scientific, Vilnius, Lithuania), 0.5 µl of each primer pair (final concentration of 10 µM of each primer, 2.5 µl of template DNA and 1.5 µl of nuclease-free water. The following thermal cycling conditions used: initial denaturation at 95 °C for 4 min, 30 cycles of denaturation at 95 °C for 30 s, amplification at 46.1 °C for 1 min, elongation at 72 °C for 1 min and a final extension at 72 °C for 7 min. All reactions were carried out in a T100 thermal cycler (Bio-Rad, Hercules, California, USA) [19]. All reactions included a positive control **(Table S1)** and a “no template control (NTC) “. The PCR products were electrophoresed at 90 V on a 1.8% gel run in Tris-borate-EDTA (0.5X) containing 0.5 µg/ml ethidium bromide and visualised using the Gel Doc XR+ imaging system (Bio-Rad, Hercules, California, USA). Of the 620 samples taken, 295 *Enterococcus* spp. were obtained, of which 245 were confirmed as *Enterococcus faecalis* via phenotypic and molecular assays. A sub-sample of all the 38 vancomycin-intermediate *E. faecalis* isolates were selected for the genotypic characterization by WGS and bioinformatics analysis (Table 1).

**Table 1:** Summary of the hospital levels, the source of sample collected, sample type, and genotypic characteristics of the *E. Faecalis* isolates

| Isolate ID | Sample Details |  |  | Typing | Antibiotic resistance genes (ARGs) |
| --- | --- | --- | --- | --- | --- |
|  | Hospital | Source | Ward | MLST (15) |  |
| 1MPA1 | Central | PHONE | PEAD | ST40 | *tetM, mphD, Isa(A)* |
| 1MPA3 | Central | PHONE | PEAD | ST40 | *tetM, mphD, Isa(A)* |
| 1MPD4 | Central | PATIENT FILE | PEAD | ST16 | *ermB, tetM, mphD, Isa(A), catA, dfrG, dfrK, Str* |
| 1MPF1 | Central | MOP | PEAD | ST498 | *mphD, Isa(A)* |
| 1MPF3 | Central | MOP | PEAD | ST498-LIKE | *tetM, mphD, Isa(A)* |
| 1MPJ101 | Central | OCCUPIED BED | PEAD | ST40 | *tetM, mphD, Isa(A)* |
| 1MPK2 | Central | NURSES TABLE | PEAD | ST40 | *tetM, mphD, Isa(A)* |
| 1MPK3 | Central | NURSES TABLE | PEAD | ST40 | *tetM, mphD, Isa(A)* |
| 1MPK4 | Central | NURSES TABLE | PEAD | ST40 | *tetM, mphD, Isa(A)* |
| 2MPJ104 | Central | OCCUPIED BED | PEAD | ST23-LIKE | *tetM, mphD, Isa(A)* |
| 3MPH1 | Central | SINK | PEAD | ST610 | *ermB, tetM, mphD, Isa(A), tetL* |
| 3MPJ101 | Central | OCCUPIED BED | PEAD | ST258 | *mphD, Isa(A)* |
| 2UIJ104 | Tertiary | OCCUPIED BED | ICU | ST126 | *tetM, mphD, Isa(A)* |
| 2UIK2 | Tertiary | NURSES TABLE | ICU | ST126 | *tetM, mphD, Isa(A)* |
| 2UIK3 | Tertiary | NURSES TABLE | ICU | ST21 | *ermB, tetM, mphD, Isa(A)* |
| 2UPA3 | Tertiary | PHONE | PEAD | ST386-LIKE | *mphD, Isa(A)* |
| 2UPC4 | Tertiary | BP APPARATUS | PEAD | ST386-LIKE | *mphD, Isa(A)* |
| 2UPF4 | Tertiary | MOP | PEAD | ST314 | *mphD, Isa(A)* |
| 2UPJ202 | Tertiary | UNOCCUPIED BED | PEAD | ST386-LIKE | *mphD, Isa(A)* |
| 3UIA2 | Tertiary | PHONE | ICU | ST16 | *tetM, mphD, Isa(A), catA, dfrG, Str* |
| 3UIC1 | Tertiary | BP APPARATUS | ICU | ST16 | *ermB, tetM, mphD, Isa(A), catA, dfrG, sat4A, aph3-III, ant6-la, aac6-Aph2* |
| 3UIE2 | Tertiary | VENTILATOR | ICU | ST268 | *tetM, mphD, Isa(A)* |
| 3UIJ202 | Tertiary | UNOCCUPIED BED | ICU | ST282 | *ermB, -----, mphD, Isa(A), catA, dfrG, sat4A, aph3-III, ant6-la, tetL* |
| 3UPF3 | Tertiary | MOP | PEAD | ST16 | *ermB, tetM, mphD, Isa(A), dfrG* |
| 3UPF4 | Tertiary | MOP | PEAD | ST16 | *ermB, tetM, mphD, Isa(A), dfrG* |
| 3UPH1 | Tertiary | SINK | PEAD | ST23 | *tetM, mphD, Isa(A)* |
| 3UIC2 | Tertiary | BP APPARATUS | ICU | ST16 | *ermB, tetM, mphD, Isa(A), catA, dfrG, sat4A, aAph3-III, ant6-la, aac6-Aph2* |
| 1CIB1 | Regional | DRIP STAND | ICU | ST21 | *ermB, tetM, mphD, Isa(A)* |
| 1CID1 | Regional | PATIENT FILE | ICU | ST21 | *ermB, tetM, mphD, Isa(A)* |
| 1CIH3 | Regional | SINK | ICU | ST21 | *ermB, tetM, mphD, Isa(A)* |
| 1CPK2 | Regional | NURSES TABLE | PEAD | ST21 | *ermB, tetM, mphD, Isa(A)* |
| 1CPK3 | Regional | NURSES TABLE | PEAD | ST126 | *tetM, mphD, Isa(A)* |
| 2CPF3 | Regional | MOP | PEAD | ST41 | *tetM, mphD, Isa(A)* |
| 2CPH2 | Regional | NURSES TABLE | PEAD | ST16-LIKE | *ermB, tetM, mphD, Isa(A)* |
| 3CPH1 | Regional | SINK | PEAD | ST23 | *tetM, mphD, Isa(A)* |
| 2SIL2 | District | DOOR HANDLE | ICU | ST922 | *ermB, tetM, mphD, Isa(A), catA, dfrG, dfrK, sat4A, aph3-III, ant6-la, tetL, fexA, optrA* |
| 2SPJ101 | District | OCCUPIED BED | ICU | ST6 | *ermB, tetM, mphD, Isa(A), catA* |
| 2SPL2 | District | DOOR HANDLE | PEAD | ST314 | *ermB, tetM, mphD, Isa(A)* |

PEAD: Paediatric ward; ICU: Intensive care unit.

### 2.4. Antibiotic susceptibility testing (AST)

Antibiotic susceptibility of the isolates was determined using the Kirby-Bauer disk diffusion method [20] according to Clinical and Laboratory Standards Institute (CLSI) guidelines [21]. The following antibiotics were used: ampicillin (10 µg), penicillin (5 µg), vancomycin (30 µg), teicoplanin (30 µg), erythromycin (15 µg), tetracycline (30 µg), ciprofloxacin (5 µg), levofloxacin (5 µg), nitrofurantoin (300 µg), chloramphenicol (30 µg), linezolid (30 µg) and rifampicin (5 µg). All discs were sourced from Oxoid (Basingstoke, United Kingdom). *Staphylococcus aureus* ATCC 25923 was used as the control. High-level aminoglycoside resistance was determined using gentamicin (120 µg) and streptomycin (300 µg) discs on Mueller-Hinton agar (Oxoid, Hampshire, England) with *E. faecalis* ATCC 29212 as the control isolate.

### 2.5. DNA isolation, genome sequencing, assembly and annotation

Genomic DNA (gDNA) was extracted using GenElute bacterial genomic DNA kit (Sigma– Aldrich, St. Louis, Missouri, United States) according to the manufacturer ‘s instructions. The quantification of extracted gDNA was determined on a Nanodrop ND1000 spectrophotometer (Thermo Scientific, Waltham, USA), Qubit 2.0 fluorometer (Invitrogen, Oregon, USA) and verified on an agarose gel electrophoresis. Multiplexed paired-end libraries (2 × 300 bp) were prepared using the Nextera DNA Flex sample preparation kit (Illumina, San Diego, California, United States) and sequences determined on an Illumina MiSeq platform with 100× coverage at the National Institute of Communicable Diseases Sequencing Core Facility, South Africa. The resulting raw reads were checked for quality, trimmed and de novo assembled into contigs using the CLC Genomics Workbench version 10.1 (CLC, Bio-QIAGEN, Aarhus, Denmark). Default parameters were used for all software unless otherwise specified. The CheckM tool version 0.9.7 [22] was used to verify that the sequence reads were not from mixed-species using lineage-specific marker sets from other genetically well-characterised closely-related *E. faecalis* isolates. The *de-novo* assembled reads were uploaded in GenBank and annotated using National Centre for Biotechnology Information (NCBI) prokaryotic genome annotation pipeline and Rapid Annotations using Subsystems Technology (RAST) 2.0 server [23].

### 2.6. WGS-based molecular typing of *E. faecalis* isolates

Multilocus sequence typing (MLST) typing was performed in-silico using the WGS data online platform tool MLST 1.8 [24] which also predicted the allelic profiles of the seven housekeeping genes, *aroE, gdh, gki, gyd, psts, xpt, and yqil* of *E. faecalis* as described previously [25].

### 2.7. Phylogenomic analysis of *Enterococcus faecalis* isolates

The de novo-assembled contigs were uploaded, and the analysis was submitted to CSI (Call SNPs & Infer) Phylogeny-1.4 (https://cge.cbs.dtu.dk/services/CSIPhylogeny-1.2), an online service which identifies single-nucleotide polymorphism (SNPs) from WGS data, filters and validates the SNP positions, and then infers phylogeny based on concatenated SNP profiles [26]. The pipeline was run with default parameters: a minimal depth at SNP positions of 10 reads, a minimal relative depth at SNP positions of 10%, a minimal distance between SNPs of 10 bp, a minimal *Z-score* of 1.96, a minimal SNP quality of 30 and a minimal read mapping quality of 25. A bootstrapped with 100 replicates indicator was applied to identify recombined regions and provide the phylogenetic accuracy in groups with little homoplasy. The Figtree (http://tree.bio.ed.ac.uk/software/figtree/) was used to edit and visualise the phylogenetic tree. The phylogeny was visualised alongside metadata for isolate demographics (including hospital, source, ward), sequence type and antibiotic resistome using Phandango [27] to provide a comprehensive analysis of the generated phylogenomic tree.

### 2.8. Genomic identification of the antibiotic resistome and mobile genetic elements (MGEs)

The bacterial analysis pipeline, ResFinder [28] was used to annotate and identify antibiotic-resistant genes using default parameters (threshold ID of 90% and a minimum length of 60%). Plasmid replicons were predicted through PlasmidFinder [29] (https://cge.cbs.dtu.dk/services/PlasmidFinder/). The PHAge Search Tool (PHAST; http://phast.wishartlab.com/) [30] server was used for the identification, annotation, and visualization of prophage sequences. The assembled genomes were further analysed for insertion sequences and transposons using ISFinder (https://isfinder.biotoul.fr/) [31]. RAST SEEDVIEWER (https://rast.nmpdr.org/seedviewer.cgi) [32] and Integrall database (http://integrall.bio.ua.pt/) [33] was also used to annotate and identify the investigated genomes for integrons and associated gene cassettes.

### 2.9. Data availability

The raw read sequences and the assembled whole-genome contigs have been deposited in GenBank. The data is available under project number **PRJNA523601**.

## 3.0. RESULTS

### 3.1. Prevalence and Antibiotic susceptibility testing

Of the 620 isolated samples, 295 were identified as *Enterococcus* spp. viz; *E. faecalis* 245 (83.1%) and other Enterococci spp. 50 (16.9%) in all the hospitals. The *E. faecalis* distributions in the hospital were 27 (93%) from the district hospital [with samples only isolated from the phone (1), mops (6), occupied beds (5), nurse ‘s table (7) and door handles (8)], 86 (85.1%) from the regional hospital [samples were isolated from all sites, phones (8), drip-stand (1), bp apparatus (6), patient files (7), ventilators (4), mops (14), sinks (9), occupied beds (14), unoccupied beds (3) nurse ‘s table (12) and door handles (8)].

From the tertiary hospital, 86 (77.5%) [Samples were also isolated from all sites, phones (16), drip-stand (9), bp apparatus (6), patient files (8), ventilators (3), mops (12), sinks (5), occupied beds (9), unoccupied beds (7) nurse ‘s table (7) and door handles (4)]. From the central hospital, 48 (88.9%) samples were isolated from phones (7), patient files (6), mop (8), sink (4), occupied bed (12), unoccupied bed (3) and nurse ‘s table (8)]. The sites with the highest contamination rates were the occupied beds and the mops with 30.2% each. In the district hospital, most positive samples identified were from the door handles 27.6%, the nurses’ tables with 24.1% and mops with 20.6%. In the regional hospital, the mops and the occupied beds 13.8% each and the nurses tables 11.9%. For the tertiary hospital, the ward phones with 14.4% and mop 10.8%. While in the central, were the occupied beds 22.2%, followed by the nurses’ tables and mop with 20.3% each.

Antibiotic susceptibility testing revealed that none of the 245 identified *E. faecalis* isolates was vancomycin-resistant (VRE). However, a total of thirty-eight (38) were of intermediate susceptibility to vancomycin and were selected for genotypic characterization by WGS and bioinformatics analysis (Table 1). These 38 vancomycin intermediate isolates showed high resistance to both tetracycline (n=30, 79%) followed by resistance to erythromycin (n=18, 47%). A small number of the isolates showed aminoglycoside resistance (gentamicin [n=4] and streptomycin [n=6]). Majority of the isolates were susceptible to ampicillin, penicillin, teicoplanin and levofloxacin whiles all the isolates were susceptible nitrofurantoin (Table S2).

### 3.2. WGS-based species confirmation and molecular typing

The identification of *E. faecalis* isolates was confirmed with generated genomic data via the Global Platform for Genomic Surveillance (Pathogenwatch). MLST-analyses (ST) revealed that the *E. faecalis* in the provincial public health-care facilities were multiclonal belonging to 15 different STs with the 6 major STs belonging to ST16 (n =7), ST40 (n = 6), ST21 (n =5), ST126 (n = 3), ST23 (n =3) and ST386 (n=3) (Table 1), with diverse allelic profiles. Moreover, one isolate belonged newly defined ST bearing a novel allele (ST922) [34].

### 3.3. Antibiotic resistance genes of *E. Faecalis* isolates

In total, 14 antibiotic resistance genes (ARGs) and variants were detected (Table 1). There were no specific differences in the resistome with regards to their hospital levels and wards. The frequency of ARGs ranged between 2–13 genes, with fifteen isolates carrying 3 resistance genes. Acquired ARGs conferring resistance to tetracycline [*tet(M)* and *tet(L)*], macrolide-lincosamide-streptogramin B (MLS_B_) [*erm(B)* and *mphD*], aminoglycosides (*sat4A, aph3-lll, ant6-la, aac6-aph2*), trimethoprim-sulfamethoxazole (*dfrG* and *dfrK*) and phenicols (*catA* and *optrA*) were found in the isolates as shown in Table 1. The *tet(M)* and *erm(B)* genes were found in 82% (31/38) and 42% (16/38) of the isolates, respectively. The *dfrG* gene predominately caused resistance to trimethoprim-sulfamethoxazole (Table 1 and Table S2).

### 3.4. WGS detection of mobile genetic support

WGS analysis revealed 11 different plasmid replicons from seven *rep* families that occurred in different combinations in the *E. faecalis* isolates (Table 2). pTEF2 (rep9), pTEF3 (repUS13), pAD1 (rep9) and pEFC1 (rep6) were the most predominant replicon types occurring in 14 (37%), 13 (34%), 13 (34%) and 9 (24%) isolates, respectively. Of note, two isolates 2SIL2 and 2SPJ101 from hospital D concomitantly harboured unique plasmid replicons (pk214 (rep7), pEFR (rep11), pPD1 (rep9), pRE25 (rep2), pUB110 (repUS14), pKH7 (rep7)) that were absent in the other isolates (Table 2). Eight (21%) of the isolates did not possess any plasmid replicons. The replicons harboured by the isolates were clonally related. For instance, major replicon pTEF2 (rep9) was harboured by isolates belonging to ST21 while the replicon set pTEF3 (repUS13), pAD1 (rep9) and pEFC1 (rep6) were harboured in ST40 isolates. Furthermore, most of the isolates (n=5) belonging to ST16 lacked plasmids.

**Table 2:**
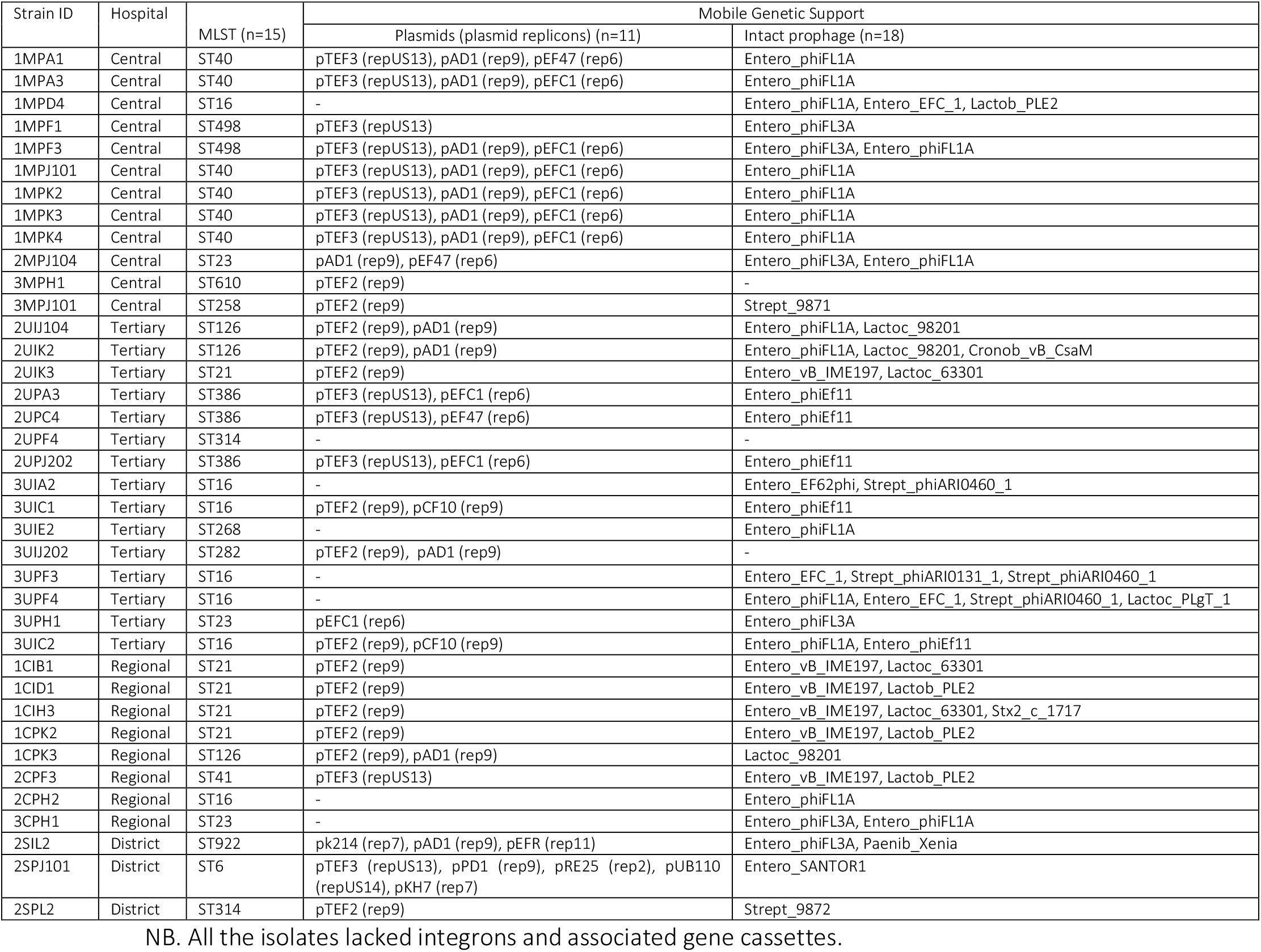
Genomic analysis of mobile genetic elements (MGEs).

| Strain ID | Hospital | MLST (n=15) | Mobile Genetic Support |  |
| --- | --- | --- | --- | --- |
|  |  |  | Plasmids (plasmid replicons) (n=11) | Intact prophage (n=18) |
| 1MPA1 | Central | ST40 | pTEF3 (repUS13), pAD1 (rep9), pEF47 (rep6) | Entero_phiFL1A |
| 1MPA3 | Central | ST40 | pTEF3 (repUS13), pAD1 (rep9), pEFC1 (rep6) | Entero_phiFL1A |
| 1MPD4 | Central | ST16 | - | Entero_phiFL1A, Entero_EFC_1, Lactob_PLE2 |
| 1MPF1 | Central | ST498 | pTEF3 (repUS13) | Entero_phiFL3A |
| 1MPF3 | Central | ST498 | pTEF3 (repUS13), pAD1 (rep9), pEFC1 (rep6) | Entero_phiFL3A, Entero_phiFL1A |
| 1MPJ101 | Central | ST40 | pTEF3 (repUS13), pAD1 (rep9), pEFC1 (rep6) | Entero_phiFL1A |
| 1MPK2 | Central | ST40 | pTEF3 (repUS13), pAD1 (rep9), pEFC1 (rep6) | Entero_phiFL1A |
| 1MPK3 | Central | ST40 | pTEF3 (repUS13), pAD1 (rep9), pEFC1 (rep6) | Entero_phiFL1A |
| 1MPK4 | Central | ST40 | pTEF3 (repUS13), pAD1 (rep9), pEFC1 (rep6) | Entero_phiFL1A |
| 2MPJ104 | Central | ST23 | pAD1 (rep9), pEF47 (rep6) | Entero_phiFL3A, Entero_phiFL1A |
| 3MPH1 | Central | ST610 | pTEF2 (rep9) | - |
| 3MPJ101 | Central | ST258 | pTEF2 (rep9) | Strept_9871 |
| 2UIJ104 | Tertiary | ST126 | pTEF2 (rep9), pAD1 (rep9) | Entero_phiFL1A, Lactoc_98201 |
| 2UIK2 | Tertiary | ST126 | pTEF2 (rep9), pAD1 (rep9) | Entero_phiFL1A, Lactoc_98201, Cronob_vB_CsaM |
| 2UIK3 | Tertiary | ST21 | pTEF2 (rep9) | Entero_vB_IME197, Lactoc_63301 |
| 2UPA3 | Tertiary | ST386 | pTEF3 (repUS13), pEFC1 (rep6) | Entero_phiEf11 |
| 2UPC4 | Tertiary | ST386 | pTEF3 (repUS13), pEF47 (rep6) | Entero_phiEf11 |
| 2UPF4 | Tertiary | ST314 | - | - |
| 2UPJ202 | Tertiary | ST386 | pTEF3 (repUS13), pEFC1 (rep6) | Entero_phiEf11 |
| 3UIA2 | Tertiary | ST16 | - | Entero_EF62phi, Strept_phiARI0460_1 |
| 3UIC1 | Tertiary | ST16 | pTEF2 (rep9), pCF10 (rep9) | Entero_phiEf11 |
| 3UIE2 | Tertiary | ST268 | - | Entero_phiFL1A |
| 3UIJ202 | Tertiary | ST282 | pTEF2 (rep9), pAD1 (rep9) | - |
| 3UPF3 | Tertiary | ST16 | - | Entero_EFC_1, Strept_phiARI0131_1, Strept_phiARI0460_1 |
| 3UPF4 | Tertiary | ST16 | - | Entero_phiFL1A, Entero_EFC_1, Strept_phiARI0460_1, Lactoc_PLgT_1 |
| 3UPH1 | Tertiary | ST23 | pEFC1 (rep6) | Entero_phiFL3A |
| 3UIC2 | Tertiary | ST16 | pTEF2 (rep9), pCF10 (rep9) | Entero_phiFL1A, Entero_phiEf11 |
| 1CIB1 | Regional | ST21 | pTEF2 (rep9) | Entero_vB_IME197, Lactoc_63301 |
| 1CID1 | Regional | ST21 | pTEF2 (rep9) | Entero_vB_IME197, Lactob_PLE2 |
| 1CIH3 | Regional | ST21 | pTEF2 (rep9) | Entero_vB_IME197, Lactoc_63301, Stx2_c_1717 |
| 1CPK2 | Regional | ST21 | pTEF2 (rep9) | Entero_vB_IME197, Lactob_PLE2 |
| 1CPK3 | Regional | ST126 | pTEF2 (rep9), pAD1 (rep9) | Lactoc_98201 |
| 2CPF3 | Regional | ST41 | pTEF3 (repUS13) | Entero_vB_IME197, Lactob_PLE2 |
| 2CPH2 | Regional | ST16 | - | Entero_phiFL1A |
| 3CPH1 | Regional | ST23 | - | Entero_phiFL3A, Entero_phiFL1A |
| 2SIL2 | District | ST922 | pk214 (rep7), pAD1 (rep9), pEFR (rep11) | Entero_phiFL3A, Paenib_Xenia |
| 2SPJ101 | District | ST6 | pTEF3 (repUS13), pPD1 (rep9), pRE25 (rep2), pUB110 (repUS14), pKH7 (rep7) | Entero_SANTOR1 |
| 2SPL2 | District | ST314 | pTEF2 (rep9) | Strept_9872 |

NB. All the isolates lacked integrons and associated gene cassettes.

The prophage analysis revealed all isolates hosted at least one intact bacteriophage except for three isolates belonging to different STs (Table 2). The predominant intact bacteriophages found were the Entero_phiFL1A (n=16, 42%), Entero_phiFL3A (n=6, 16%), Entero_vB_IME197 (n=6, 16%) and Entero_phiEf11 (n=5, 13%). Four prophages were identified in one *E. faecalis* ST16 (3UPF4) strain isolated from the mop of a paediatric ward in hospital B with a peculiar bacteriophage (Lactoc_PLgT_1). The isolates 1C1H3, 1MPD4, 2U1K2 and 2UPF3 from different hospitals hosted 3 prophages. The prophage harboured by the isolates were clonally related (Table 2).

A myriad of IS families was found in the isolates with no association with respect to the hospital and ward. The 5 major IS families were IS3 (predicted to be linked with *Enterococcus faecium/Streptococcus agalactiae* sources), IS5 (predicted to be associated with *Cyanotheca sp*. sources), IS1595 (predicted to be linked with *Bacillus subtilis*), ISL3 (predicted to be linked with *Streptococcus mutans/thermophilus*) and IS607 (predicted to be linked with both *Campylobacter sp*. and *Virus NY2A*), (Table S3). The transposase (Tn*3*) linked with *Bacillus thuringiensis* was found in 7 of the isolates identified from different sources (Table S3). All the isolates lacked integrons and their associated gene cassettes.

### 3.5. Phylogenomic and metadata analysis

A phylogenetic tree reconstructed to analyse genetic relationships between the isolates revealed a high divergence of isolates according to the different levels of care (Figure 1). For instance, each hospital was generally associated with specific dominant clones (i.e., ST40 and ST498 were mostly found in hospital A; ST16, ST126, and ST386 were found in hospital B; and ST21 was predominately found in hospital C (Table 1 and Figure 1).

**Figure 1:**
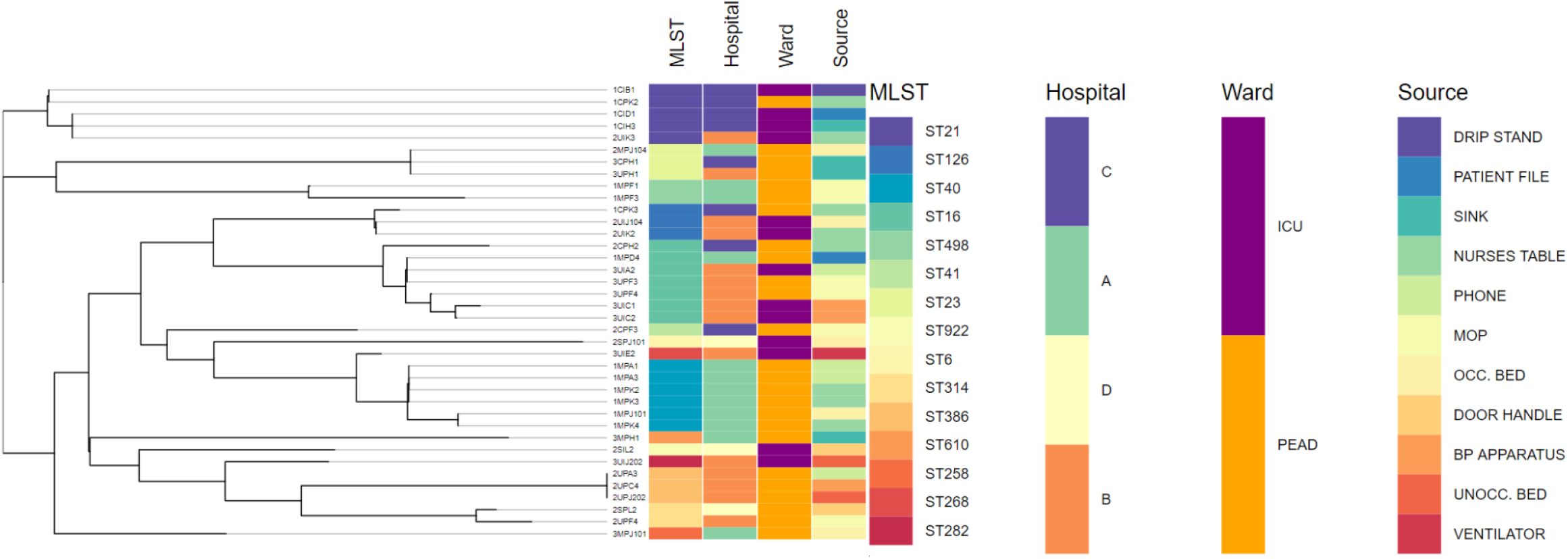
The whole genome MLST phylogenomic branch and metadata of isolate information (including isolate identity, hospital, source and ward) and WGS *in-silico typing* (sequence type and antibiotic resistome) coupled using Phandango (https://jameshadfield.github.io/phandango/#). *E. faecalis* isolates at different level of care in Durban, South Africa. The linking lines in the phylogenetic tree differentiate between the different clades. Metadata annotations show that there were generally distinct major sequence types between the 4 hospital environments however within each hospital there was the spread of these major clones between different sources in the wards.

Phylogenomic trees coupled metadata visualization analysis provided a more in-depth insight into the characteristics and distinctions between isolates and revealed the intra-clonal spread of *E. faecalis* strains between different sources within the same hospitals (Figure 1).

Specifically, ST21 was found on the drip stand, patient file, sink and nurses table in both ICU and paediatric ward of hospital C. Similarly, ST40 was found on phone, patient file, mop, occupied bed and nurses table of the paediatric ward hospital A. The ST16 clone was isolated on the mop (paediatric ward), phone and BP apparatus (ICU) of hospital B. More so, ST386 linked with the phone, BP apparatus, and unoccupied bed in the paediatric ward of hospital B while ST126 was found on the occupied bed and nurses table in the ICU of the same hospital.

## 4.0. Discussion

In line with the global trend, reports on bacterial contamination in hospital environments is increasing in Africa across all sectors [35], and *E. faecalis* is one of the most common enterococcal species isolated from the hospital environment. This is evident from our results where *E. faecalis* (n=245) was the most prevalent organisms compared to *E. faecium* (n= 53). *E. faecalis* is recognised as an important hospital-associated pathogen responsible for approximately 80-90% of cases reported in the hospital settings followed by 5-10% *E. faecium* [36] and hence has been placed in the category of pathogens posing a major threat to healthcare systems [37]. Furthermore, *E. faecalis* represent major infection prevention attributed to their ability to persist for long periods on hands and remain viable on environmental surfaces (inanimate surfaces) due to their microbial structure thus, can serve as a reservoir for ongoing transmission in the absence of regular decontamination [38]. Additionally, *E. faecalis* posses the ability to acquire additional resistance through the transfer of mobile genetic support such as plasmids, prophages, and insertion sequences [39,40]. The acquisition of resistance and genetic support poses a therapeutic challenge.

The WGS results showed that none of the *E. faecalis* harbored vancomycin-resistant genes which corroborated with the phenotypes [12]. This affirms the view of Ellington *et al* on the role of WGS in antimicrobial susceptibility testing of bacteria for the explanation of phenotypic result for samples [41] and further confirms WGS as a more discriminatory tool to infer antibiotic susceptibility as compared to relying fully on phenotypic testing alone. Majority of the isolates were susceptible to ampicillin, penicillin, teicoplanin, levofloxacin, and nitrofurantoin confirming their use as treatment options in South Africa, particularly ampicillin (the drug of choice for *E. faecalis* infections) [42].

Tetracycline demonstrated reduced susceptibility against *E. faecalis* mediated mostly by the ribosomal protection protein, *tet(M)* [11,43]. This was consistent with previous studies that found the *tet(M)* as the dominant gene causing tetracycline resistance in *E. faecalis* isolates across all the one-health sectors (human-animal-environment interface) [35]. For instance, in a 2014 hospital-based study in China by Jia *et al*.[44], tet*(M)* was found to cause tetracycline-resistant *E. faecalis* isolates. Similarly, *Said et al*. [45] also detected tet*(M)* as 96.1% of all tetracycline-resistant *Enterococcus* isolates in Egypt. However, the tetracycline resistance exhibited by 2SIL2 isolate was mediated by both ribosomal-protection gene [*tet(M)*] and active-efflux gene [*tet(L)*]. This indicates the significant role played by efflux pumps in mediating antibiotic resistance [46]. The low prevalence of the tet*(L)* was not unusual and pointed to the fact that ribosomal protection protein is the main mechanism of tetracycline-resistant *E. faecalis* isolates. The moderate level of erythromycin resistance was mediated by *erm(B)* genes which are the most common mechanism of resistance reported for the macrolide class of antibiotics in Africa [35] and globally [11,47] for *Enterococcus*. There was small number of isolates showing aminoglycoside resistance across the different levels of care, which corresponded to the aminoglycoside-modifying enzymes found. However, these isolates exhibited high-level resistance encoding a set of enzymes (*sat4A, aph3-lll, ant6-la, aac6-aph2*). However, this was not unusual as some *Enterococcus* spp. are known to produce low-level resistance to aminoglycosides by limiting drug uptake, which is associated with the proteins involved in electron transport [48]. More so, the *OptrA* gene implicated in linezolid resistance was found in only one isolate (2SIL2) however, it was unexpressed as the isolate was susceptible to linezolid (Table 1 and S2) as reported in study in China were *OptrA* was found in eighteen linezolid non-resistant enterococci [49].

A noticeable polyclonal nature was observed in the *E. faecalis* isolates with 15 distinct STs, including one novel STs, highlighting the diverse nature of the strains in the province. The major STs found such as ST16, ST40 and ST21 were previously reported in Saudi Arabia, China, Tunisia, France, and Spain from human subjects, hospitalised patients, animals and wastewater [36,50–53]. Similarly, other studies have also reported the ST126, ST23 and ST386 in different settings (human, animal and environment) and hence do not suggest any kind of host specificity in these major STs reported in this study [54]. However, unlike other countries, the population structure of *E. faecalis* from different settings in South Africa are minimally monitored, if at all, making it difficult to correlate our results with studies in South Africa. This calls for the need for *E. faecalis* to be included in surveillance schemes to enable the monitoring of the molecular epidemiology of isolates collected over larger tempo-spatial scales using high throughput technologies such as WGS [55]. Such surveillance would help microbiologists and public health practitioners to gain better insights into the evolution and dissemination of *E. faecalis*.

Characterizing the genetic support in the isolates indicated that the majority of *E. faecalis* in the different hospitals are likely reservoirs for diverse mobile genetic elements and associated ARGs (especially for tetracycline, erythromycin). There was a higher plasmid prevalence rate (seven *rep* families) and the detection of two or more distinct replicons in one strain. Accordingly, this finding agrees with the fact that numerous types of plasmids are often present in enterococci from a clinical setting [56–58]. More so, other studies have shown that single isolates of *E. faecalis* may harbour multiple plasmids [57,59].

There was no specific pattern between the acquisition of insertion sequence families or transposable elements with respect to the ward and level of care however the presence of major IS families in *E. faecalis* clones imply that these elements are spread by horizontal gene transfer (HGT) [39]. Moreover, the acquisition of these elements can lead to transposition in the genome aid in the transfer of resistance genes, enabling it to adapt to new environmental challenges and colonise new niches [60]. For instance, IS3 family upstream of the *EmrB* gene has been reported for enhanced erythromycin resistance [60]. The ability of these clones to acquire novel genetic features may contribute to their increased persistence and highlights its potential public health threat.

Comparative phylogenomics using WGS SNPs analysis revealed a higher genetic diversity between the strains with respect to each specific hospital. This implied that the major clones were mostly hospital-specific, which was in concordance with the *in-silico* MLST typing scheme (Figure 1). Interestingly, a study by Kawalec *et al*., [61] also found a higher diversity in the clonal structure of *E. faecalis* strains among hospitals in Poland. Visualizing the phylogenomic tree with metadata revealed the major clones in the various hospitals. This further depicted the intra-clonal spread of *E. faecalis* strains between different sources within the same hospital, reiterating the need for phylo/meta-analysis to increase confidence in molecular epidemiological studies. For instance, at the paediatric ward of hospital A, the ST40 clone was isolated from a phone, nurses table, patient file, mop and occupied bed which may be due to hand contamination by patients and/or healthcare workers (nurses, janitor staff, etc.) (Figure 1). A similar scenario occurred in hospital B, where ST386 was found in the paediatric ward (on the phone, BP apparatus and unoccupied bed) while the ST126 was isolated in the ICU (on nurses table and occupied bed). Reports on enterococci transient carriage on the hands of healthcare workers and patients as well their presence on, medical equipment, or environmental surfaces has been documented in several studies [62–65]. Other studies have reported the movement of colonised patients among different settings in the hospital as responsible for these patterns of transmission [64,66]. Moreover, hospitals B and C observed intra-ward spreads (both ICU and paediatric ward) of ST16 and 21, respectively from different sites with each hospital. The transmission of enterococcal strains has been documented within medical units, given credence to the study findings [67,68].

Frequent contact with healthcare providers and movement of colonised patients among different healthcare settings is a probable means for these patterns of transmission in hospitals A, B and C. However, there were limited isolates from district hospital (Hospital D) due to the number of isolates obtained for any detailed comparative analysis. Even though the findings of our study may not be generalised to the overall situation in the country, this study improves the understanding of the prevalence, genetic content, and relatedness of *E. faecalis* contamination of hospital environments. It is thus recommended that scheduled periodic identification of transmitting sources in the hospitals’ inanimate environment, strict enforcement and adhesions of IPC practices amongst the health workers and isolation of colonised patients should be imposed to reduce the incidence and transmission of *E. faecalis* hospital environments. More so, the study was limited by the number of isolates selected for sequencing and hence there is the need for large-scale genomic epidemiology to elucidate the population structure in the various hospital environments in South Africa.

## 5.0. Conclusion

This genomic analysis provided a snapshot of the hospital inanimate environment as a reservoir of resistant *E. faecalis*, its associated mobilome (plasmids, prophages, insertion sequences and transposons) and revealed a complex intra-clonal spread of *E. faecalis* major clones between the sites within each specific hospital setting. This study improves our understanding of the dissemination of *E. faecalis* in hospital environments and will aid in the design of optimal infection prevention and control strategies in clinical settings.

## Data Availability

The generated data used to support the findings of this study are included in the article.

## Acknowledgment

We are grateful to the Sequencing Core Facility, National Institute for Communicable Diseases, National Health Laboratory Service, Johannesburg, South Africa; the South African Research Chairs Initiative of the Department of Science and Technology and the National Research Foundation of South Africa (Grant number: **98342**); and the College of Health young researcher grant, University of KwaZulu-Natal, for sponsoring this research.

## General Disclaimer

Any opinions, findings and conclusions or recommendations expressed in this material are those of the author(s) and do not necessarily reflect the views of the organisations or agencies that provided support for the project. The funders had no role in the study design, nor the decision to submit the work for publication.

## Competing interests

The authors have no competing interests to declare.

## Author Contributions

**C.S** co-conceptualised the study, undertook sample collection, microbiological and bioinformatic analyses and drafted the manuscript. **D.G.A**. co-conceptualised the study, undertook bioinformatics analyses, data interpretation and a critical revision of the manuscript. **M.A** and **A.I**. performed whole-genome sequencing analysis and critical revision of the manuscript. **S.Y.E**. and **L.B** co-conceptualised the study, supervised the study and undertook critical revision of the manuscript.

## Supplemental Material

Table S1: List of genus-and species-specific primers and control strains used in this study. Table S2: Antibiotic susceptibility profiles of *E. faecalis* (n=38).

Table S3: Distribution of 6 major insertion sequence (IS)/transposase families and their associated predicted sources among *E. faecalis* isolates via the ISFinder database.

---

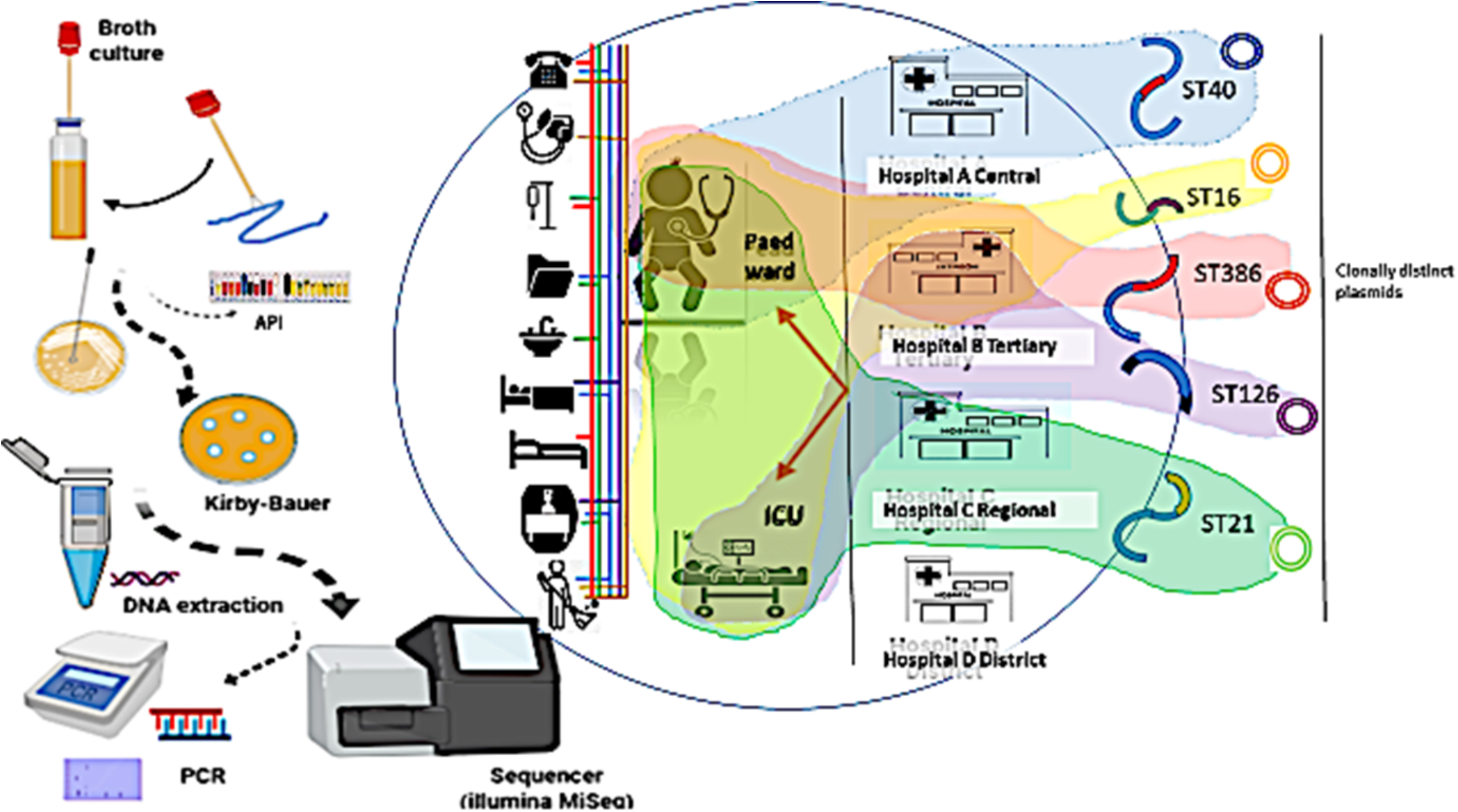

## Notes

### Competing Interest Statement

The authors have declared no competing interest.

## References

1. Comar, M.; D’Accolti, M.; Cason, C.; Soffritti, I.; Campisciano, G.; Lanzoni, L.; Bisi, M.; Volta, A.; Mazzacane, S.; Caselli, E. Introduction of NGS in Environmental Surveillance for Healthcare-Associated Infection Control. Microorganisms 2019, 7, 708, doi:10.3390/microorganisms7120708.

2. Gilchrist, C.A.; Turner, S.D.; Riley, M.F.; Petri, W.A.; Hewlett, E.L. Whole-Genome Sequencing in Outbreak Analysis. Clin. Microbiol. Rev. 2015, 28, 541–563, doi:10.1128/CMR.00075-13.

3. Bani-Yaghoub, M.; Gautam, R.; Döpfer, D.; Kaspar, C.W.; Ivanek, R. Effectiveness of environmental decontamination as an infection control measure. Epidemiol. Infect. 2012, 140, 542–553, doi:10.1017/S0950268811000604.

4. Dusé, A. Infection control in developing countries with particular emphasis on South Africa. South Afr J Epidemiol Infect 2005, 20, 37–41.

5. Harbarth, S.; Balkhy, H.H.; Goossens, H.; Jarlier, V.; Kluytmans, J.; Laxminarayan, R.; Saam, M.; Van Belkum, A.; Pittet, D. Antimicrobial resistance: one world, one fight! Antimicrob. Resist. Infect. Control 2015, 4, 49, doi:10.1186/s13756-015-0091-2.

6. Mcgann, P.; Bunin, J.L.; Snesrud, E.; Singh, S.; Maybank, R.; Ong, A.C.; Kwak, Y.I.; Seronello, S.; Clifford, R.J.; Hinkle, M.; et al. Real time application of whole genome sequencing for outbreak investigation – What is an achievable turnaround time ? Diagn. Microbiol. Infect. Dis. 2016, 85, 277–282, doi:10.1016/j.diagmicrobio.2016.04.020.

7. Quainoo, S.; Coolen, J.P.M.; van Hijum, S.A.F.T.; Huynen, M.A.; Melchers, W.J.G.; van Schaik, W.; Wertheim, H.F.L. Whole-Genome Sequencing of Bacterial Pathogens: the Future of Nosocomial Outbreak Analysis. Clin. Microbiol. Rev. 2017, 30, 1015–1063, doi:10.1128/CMR.00016-17.

8. Balloux, F.; Brønstad Brynildsrud, O.; van Dorp, L.; Shaw, L.P.; Chen, H.; Harris, K.A.; Wang, H.; Eldholm, V. From Theory to Practice: Translating Whole-Genome Sequencing (WGS) into the Clinic. Trends Microbiol. 2018, 26, 1035–1048, doi:10.1016/j.tim.2018.08.004.

9. Zaheer, R.; Cook, S.R.; Barbieri, R.; Goji, N.; Cameron, A.; Petkau, A.; Polo, R.O.; Tymensen, L.; Stamm, C.; Song, J.; et al. Surveillance of Enterococcus spp. reveals distinct species and antimicrobial resistance diversity across a One-Health continuum. Sci. Rep. 2020, 10, 3937, doi:10.1038/s41598-020-61002-5.

10. Hollenbeck, B.L.; Rice, L.B. Intrinsic and acquired resistance mechanisms in enterococcus. Virulence 2012, 3, 421–569, doi:10.4161/viru.21282.

11. Miller, W.R.; Munita, J.M.; Arias, C.A. Mechanisms of antibiotic resistance in enterococci. Expert Rev. Anti. Infect. Ther. 2014, 12, 1221–1236, doi:10.1586/14787210.2014.956092.

12. Shobo, C.O.; Essack, S.Y.; Bester, L.A. Enterococcal contamination of hospital environments in KwaZulu-Natal, South Africa. J. Appl. Microbiol. 2021, doi:10.1111/jam.15224.

13. Iweriebor, B.; Gaqavu, S.; Obi, L.; Nwodo, U.; Okoh, A. Antibiotic Susceptibilities of Enterococcus Species Isolated from Hospital and Domestic Wastewater Effluents in Alice, Eastern Cape Province of South Africa. Int. J. Environ. Res. Public Health 2015, 12, 4231–4246, doi:10.3390/ijerph120404231.

14. Ekwanzala, M.D.; Dewar, J.B.; Kamika, I.; Momba, M.N.B. Comparative genomics of vancomycin-resistant Enterococcus spp. revealed common resistome determinants from hospital wastewater to aquatic environments. Sci. Total Environ. 2020, 719, 137275, doi:10.1016/j.scitotenv.2020.137275.

15. Gregersen, T. Rapid method for distinction of gram-negative from gram-positive bacteria. Eur. J. Appl. Microbiol. Biotechnol. 1978, 5, 123–127, doi:10.1007/BF00498806.

16. Englen, M.D.; Kelley, L.C. A rapid DNA isolation procedure for the identification of Campylobacter jejuni by the polymerase chain reaction. Lett. Appl. Microbiol. 2000, 31, 421–426, doi:10.1046/j.1365-2672.2000.00841.x.

17. Jackson, C.R.; Fedorka-Cray, P.J.; Barrett, J.B. Use of a genus-and species-specific multiplex PCR for identification of enterococci. J. Clin. Microbiol. 2004, 42, 3558–3565, doi:10.1128/JCM.42.8.3558-3565.2004.

18. Ke, D.; Picard, F.J.; Martineau, F.; Ménard, C.; Roy, P.H.; Ouellette, M.; Bergeron, M.G. Development of a PCR assay for rapid detection of enterococci. J. Clin. Microbiol. 1999, 37, 3497–503.

19. Molechan, C.; Amoako, D.G.; Abia, A.L.K.; Somboro, A.M.; Bester, L.A.; Essack, S.Y. Molecular epidemiology of antibiotic-resistant Enterococcus spp. from the farm-to-fork continuum in intensive poultry production in KwaZulu-Natal, South Africa. Sci. Total Environ. 2019, 692, 868–878, doi:10.1016/j.scitotenv.2019.07.324.

20. Yang, Y.; Jiang, X.-T.; Zhang, T. Evaluation of a hybrid approach using UBLAST and BLASTX for metagenomic sequences annotation of specific functional genes. PLoS One 2014, 9, e110947, doi:10.1371/journal.pone.0110947.

21. CLSI Performance standards for antimicrobial susceptibility testing. CLSI supplement M100.; 27th ed.; Clinical and Laboratory Standards Institute; Wayne, Pennsylvania, 2017;

22. Parks, D.H.; Imelfort, M.; Skennerton, C.T.; Hugenholtz, P.; Tyson, G.W. CheckM: assessing the quality of microbial genomes recovered from isolates, single cells, and metagenomes. Genome Res. 2015, 25, 1043–1055, doi:10.1101/gr.186072.114.

23. Aziz, R.K.; Bartels, D.; Best, A.; DeJongh, M.; Disz, T.; Edwards, R.A.; Formsma, K.; Gerdes, S.; Glass, E.M.; Kubal, M.; et al. The RAST Server: Rapid annotations using subsystems technology. BMC Genomics 2008, 9, 1–15, doi:10.1186/1471-2164-9-75.

24. Larsen, M. V; Cosentino, S.; Rasmussen, S.; Friis, C.; Hasman, H.; Marvig, R.L.; Jelsbak, L.; Sicheritz-Pontén, T.; Ussery, D.W.; Aarestrup, F.M.; et al. Multilocus Sequence Typing of Total-Genome-Sequenced Bacteria. J. Clin. Microbiol. 2012, 50, 1355–1361, doi:10.1128/jcm.06094-11.

25. Feil, E.J.; Cooper, J.E.; Grundmann, H.; Robinson, D.A.; Enright, M.C.; Berendt, T.; Peacock, S.J.; Smith, J.M.; Murphy, M.; Spratt, B.G.; et al. How Clonal Is Staphylococcus aureus? 2003, 185, 3307–3316, doi:10.1128/JB.185.11.3307.

26. Ahrenfeldt, J.; Skaarup, C.; Hasman, H.; Pedersen, A.G.; Aarestrup, F.M.; Lund, O. Bacterial whole genome-based phylogeny: construction of a new benchmarking dataset and assessment of some existing methods. BMC Genomics 2017, 18, 19, doi:10.1186/s12864-016-3407-6.

27. Hadfield, J.; Croucher, N.J.; Goater, R.J.; Abudahab, K.; Aanensen, D.M.; Harris, S.R. Phandango: an interactive viewer for bacterial population genomics. Bioinformatics 2017, 34, 292–293, doi:10.1093/bioinformatics/btx610.

28. Zankari, E.; Hasman, H.; Cosentino, S.; Vestergaard, M.; Rasmussen, S.; Lund, O.; Aarestrup, F.M.; Larsen, M.V. Identification of acquired antimicrobial resistance genes. J. Antimicrob. Chemother. 2012, 67, 2640–2644, doi:10.1093/jac/dks261.

29. Carattoli, A.; Zankari, E.; García-Fernández, A.; Voldby Larsen, M.; Lund, O.; Villa, L.; Møller Aarestrup, F.; Hasman, H. In Silico Detection and Typing of Plasmids using PlasmidFinder and Plasmid Multilocus Sequence Typing. Antimicrob. Agents Chemother. 2014, 58, 3895–3903, doi:10.1128/AAC.02412-14.

30. Zhou, Y.; Liang, Y.; Lynch, K.H.; Dennis, J.J.; Wishart, D.S. PHAST: A Fast Phage Search Tool. Nucleic Acids Res. 2011, 39, 347–352, doi:10.1093/nar/gkr485.

31. Siguier, P. ISfinder: the reference centre for bacterial insertion sequences. Nucleic Acids Res. 2006, 34, D32–D36, doi:10.1093/nar/gkj014.

32. Overbeek, R.; Olson, R.; Pusch, G.D.; Olsen, G.J.; Davis, J.J.; Disz, T.; Edwards, R.A.; Gerdes, S.; Parrello, B.; Shukla, M.; et al. The SEED and the Rapid Annotation of microbial genomes using Subsystems Technology (RAST). Nucleic Acids Res. 2014, 42, D206–D214, doi:10.1093/nar/gkt1226.

33. Moura, A.; Soares, M.; Pereira, C.; Leitão, N.; Henriques, I.; Correia, A. INTEGRALL: A database and search engine for integrons, integrases and gene cassettes. Bioinformatics 2009, 25, 1096–1098, doi:10.1093/bioinformatics/btp105.

34. Shobo, C.O.; Amoako, D.G.; Allam, M.; Ismail, A.; Essack, S.Y.; Bester, L.A. Genome Sequence of a Novel Enterococcus faecalis Sequence Type 922 Strain Isolated from a Door Handle in the Intensive Care Unit of a District Hospital in Durban, South Africa. Microbiol. Resour. Announc. 2019, 8, 1–2, doi:10.1128/MRA.00582-19.

35. Osei Sekyere, J.; Mensah, E. Molecular epidemiology and mechanisms of antibiotic resistance in Enterococcus spp., Staphylococcus spp., and Streptococcus spp. in Africa: a systematic review from a One Health perspective. Ann. N. Y. Acad. Sci. 2020, 1465, 29–58, doi:10.1111/nyas.14254.

36. Farman, M.; Yasir, M.; Al-Hindi, R.R.; Farraj, S.A.; Jiman-Fatani, A.A.; Alawi, M.; Azhar, E.I. Genomic analysis of multidrug-resistant clinical Enterococcus faecalis isolates for antimicrobial resistance genes and virulence factors from the western region of Saudi Arabia. Antimicrob. Resist. Infect. Control 2019, 8, 55, doi:10.1186/s13756-019-0508-4.

37. Zarb, P.; Coignard, B.; Griskeviciene, J.; Muller, A.; Vankerckhoven, V.; Weist, K.; Goossens, M.; Vaerenberg, S.; Hopkins, S.; Catry, B.; et al. The European Centre for Disease Prevention and Control (ECDC) pilot point prevalence survey of healthcare-associated infections and antimicrobial use. Euro Surveill. 2012, 17, 20316, doi:10.2807/ese.17.46.20316-en.

38. Kramer, A.; Schwebke, I.; Kampf, G. How long do nosocomial pathogens persist on inanimate surfaces? A systematic review. BMC Infect. Dis. 2006, 6, 130, doi:10.1186/1471-2334-6-130.

39. Mikalsen, T.; Pedersen, T.; Willems, R.; Coque, T.M.; Werner, G.; Sadowy, E.; van Schaik, W.; Jensen, L.B.; Sundsfjord, A.; Hegstad, K. Investigating the mobilome in clinically important lineages of Enterococcus faecium and Enterococcus faecalis. BMC Genomics 2015, 16, 282, doi:10.1186/s12864-015-1407-6.

40. Partridge, S.R.; Kwong, S.M.; Firth, N.; Jensen, S.O. Mobile Genetic Elements Associated with Antimicrobial Resistance. Clin. Microbiol. Rev. 2018, 31, e00088–17, doi:10.1128/CMR.00088-17.

41. Ellington, M.J.; Ekelund, O.; Aarestrup, F.M.; Canton, R.; Doumith, M.; Giske, C.; Grundman, H.; Hasman, H.; Holden, M.T.G.; Hopkins, K.L.; et al. The role of whole genome sequencing in antimicrobial susceptibility testing of bacteria: report from the EUCAST Subcommittee. Clin. Microbiol. Infect. 2017, 23, 2–22, doi:10.1016/j.cmi.2016.11.012.

42. National Department of Health Surveillance for resistance and consumption of antibiotics in South Africa; 2018;

43. Warburton, P.J.; Amodeo, N.; Roberts, A.P. Mosaic tetracycline resistance genes encoding ribosomal protection proteins. J. Antimicrob. Chemother. 2016, 71, 3333–3339, doi:10.1093/jac/dkw304.

44. Jia, W.; Li, G.; Wang, W. Prevalence and Antimicrobial Resistance of Enterococcus Species: A Hospital-Based Study in China. Int. J. Environ. Res. Public Health 2014, 11, 3424–3442, doi:10.3390/ijerph110303424.

45. Said, H.S.; Abdelmegeed, E.S. Emergence of multidrug resistance and extensive drug resistance among enterococcal clinical isolates in Egypt. Infect. Drug Resist. 2019, 12, 1113–1125, doi:10.2147/IDR.S189341.

46. Blanco, P.; Hernando-Amado, S.; Reales-Calderon, J.; Corona, F.; Lira, F.; Alcalde-Rico, M.; Bernardini, A.; Sanchez, M.; Martinez, J. Bacterial Multidrug Efflux Pumps: Much More Than Antibiotic Resistance Determinants. Microorganisms 2016, 4, 14, doi:10.3390/microorganisms4010014.

47. Tian, Y.; Yu, H.; Wang, Z. Distribution of acquired antibiotic resistance genes among Enterococcus spp. isolated from a hospital in Baotou, China. BMC Res. Notes 2019, 12, 27, doi:10.1186/s13104-019-4064-z.

48. Shete, V.; Grover, N.; Kumar, M. Analysis of Aminoglycoside Modifying Enzyme Genes Responsible for High-Level Aminoglycoside Resistance among Enterococcal Isolates. J. Pathog. 2017, 2017, 1–5, doi:10.1155/2017/3256952.

49. Li, P.; Yang, Y.; Ding, L.; Xu, X.; Lin, D. Molecular Investigations of Linezolid Resistance in Enterococci OptrA Variants from a Hospital in Shanghai. Infect. Drug Resist. 2020, Volume 13, 2711–2716, doi:10.2147/IDR.S251490.

50. Zischka, M.; Künne, C.T.; Blom, J.; Wobser, D.; Sakιnç, T.; Schmidt-Hohagen, K.; Dabrowski, P.W.; Nitsche, A.; Hübner, J.; Hain, T.; et al. Comprehensive molecular, genomic and phenotypic analysis of a major clone of Enterococcus faecalis MLST ST40. BMC Genomics 2015, 16, 175, doi:10.1186/s12864-015-1367-x.

51. Kuch, A.; Willems, R.J.L.; Werner, G.; Coque, T.M.; Hammerum, A.M.; Sundsfjord, A.; Klare, I.; Ruiz-Garbajosa, P.; Simonsen, G.S.; van Luit-Asbroek, M.; et al. Insight into antimicrobial susceptibility and population structure of contemporary human Enterococcus faecalis isolates from Europe. J. Antimicrob. Chemother. 2012, 67, 551–558, doi:10.1093/jac/dkr544.

52. Quiñones, D.; Kobayashi, N.; Nagashima, S. Molecular Epidemiologic Analysis of Enterococcus faecalis Isolates in Cuba by Multilocus Sequence Typing. Microb. Drug Resist. 2009, 15, 287–293, doi:10.1089/mdr.2009.0028.

53. McBride, S.M.; Fischetti, V.A.; LeBlanc, D.J.; Moellering, R.C.; Gilmore, M.S. Genetic Diversity among Enterococcus faecalis. PLoS One 2007, 2, e582, doi:10.1371/journal.pone.0000582.

54. Raven, K.E.; Reuter, S.; Gouliouris, T.; Reynolds, R.; Russell, J.E.; Brown, N.M.; Török, M.E.; Parkhill, J.; Peacock, S.J. Genome-based characterization of hospital-adapted Enterococcus faecalis lineages. Nat. Microbiol. 2016, 1, 15033, doi:10.1038/nmicrobiol.2015.33.

55. Amoako, D.G.; Somboro, A.M.; Abia, A.L.K.; Allam, M.; Ismail, A.; Bester, L.; Essack, S.Y. Genomic analysis of methicillin-resistant Staphylococcus aureus isolated from poultry and occupational farm workers in Umgungundlovu District, South Africa. Sci. Total Environ. 2019, 670, 704–716, doi:10.1016/j.scitotenv.2019.03.110.

56. Zhu, W.; Murray, P.R.; Huskins, W.C.; Jernigan, J.A.; McDonald, L.C.; Clark, N.C.; Anderson, K.F.; McDougal, L.K.; Hageman, J.C.; Olsen-Rasmussen, M.; et al. Dissemination of an Enterococcus Inc18-Like vanA Plasmid Associated with Vancomycin-Resistant Staphylococcus aureus. Antimicrob. Agents Chemother. 2010, 54, 4314–4320, doi:10.1128/AAC.00185-10.

57. Garcia-Migura, L.; Sanchez-Valenzuela, A.J.; Jensen, L.B. Presence of Glycopeptide-Encoding Plasmids in Enterococcal Isolates from Food and Humans in Denmark. FOODBORNE Pathog. Dis. 2011, 8, 1191–1197, doi:10.1089/fpd.2011.0897.

58. Song, X.; Sun, J.; Mikalsen, T.; Roberts, A.P.; Sundsfjord, A. Characterisation of the Plasmidome within Enterococcus faecalis Isolated from Marginal Periodontitis Patients in Norway. PLoS One 2013, 8, e62248, doi:10.1371/journal.pone.0062248.

59. Sedgley, C.; Clewell, D.B. Bacterial plasmids in the oral and endodontic microflora. Endod. Top. 2004, 9, 37–51, doi:10.1111/j.1601-1546.2004.00077.x.

60. Vandecraen, J.; Chandler, M.; Aertsen, A.; Van Houdt, R. The impact of insertion sequences on bacterial genome plasticity and adaptability. Crit. Rev. Microbiol. 2017, 43, 709–730, doi:10.1080/1040841X.2017.1303661.

61. Kawalec, M.; Pietras, Z.; Danilowicz, E.; Jakubczak, A.; Gniadkowski, M.; Hryniewicz, W.; Willems, R.J.L. Clonal Structure of Enterococcus faecalis Isolated from Polish Hospitals: Characterization of Epidemic Clones. J. Clin. Microbiol. 2007, 45, 147–153, doi:10.1128/JCM.01704-06.

62. Tajeddin, E.; Rashidan, M.; Razaghi, M.; Javadi, S.S.S.; Sherafat, S.J.; Alebouyeh, M.; Sarbazi, M.R.; Mansouri, N.; Zali, M.R. The role of the intensive care unit environment and health-care workers in the transmission of bacteria associated with hospital acquired infections. J. Infect. Public Health 2016, 9, 13–23, doi:10.1016/j.jiph.2015.05.010.

63. Agudelo Higuita, N.I.; Huycke, M.M. Enterococcal Disease, Epidemiology, and Implications for Treatment; 2014;

64. Daniel, D.S.; Lee, S.M.; Dykes, G.A.; Rahman, S. Public Health Risks of Multiple-Drug-Resistant Enterococcus spp. in Southeast Asia. Appl. Environ. Microbiol. 2015, 81, 6090–6097, doi:10.1128/AEM.01741-15.

65. Evans Patterson, J.; Sweeney, A.H.; Simms, M.; Carley, N.; Mangi, R.; Sabetta, J.; Lyons, R.W. An Analysis of 110 Serious Enterococcal Infections Epidemiology, Antibiotic Susceptibility, and Outcome. Medicine (Baltimore). 1995, 74, 191–200, doi:10.1097/00005792-199507000-00003.

66. Jackson, S.S.; Harris, A.D.; Magder, L.S.; Stafford, K.A.; Johnson, J.K.; Miller, L.G.; Calfee, D.P.; Thom, K.A. Bacterial burden is associated with increased transmission to health care workers from patients colonized with vancomycin-resistant Enterococcus. Am. J. Infect. Control 2019, 47, 13–17, doi:10.1016/j.ajic.2018.07.011.

67. D’Agata, E.M.C.; Green, W.K.; Schulman, G.; Li, H.; Tang, Y.-W.; Schaffner, W. Vancomycin-Resistant Enterococci among Chronic Hemodialysis Patients: A Prospective Study of Acquisition. Clin. Infect. Dis. 2001, 32, 23–29, doi:10.1086/317549.

68. Lund, B.; Agvald-Ohman, C.; Hultberg, A.; Edlund, C. Frequent Transmission of Enterococcal Strains between Mechanically Ventilated Patients Treated at an Intensive Care Unit. J. Clin. Microbiol. 2002, 40, 2084–2088, doi:10.1128/JCM.40.6.2084-2088.2002

