## Supplementary Material for "A Genomic Snapshot of *Enterococcus faecalis* within Public Hospital Environments in South Africa"

**Table S1:** List of genus-and species-specific primers and control strains used in this study

| Control Strain | Primer | Primer sequence 5’-3’ | Product size (bp) | Reference |
| --- | --- | --- | --- | --- |
| *E. faecalis* ATCC 51299 | ENT1  ENT2 | TACTGACAAACCATTCATGATG  AACTTCGTCACCAACGCGAAC | 112 | (Molechan et al., 2019) |
| *E. faecalis* ATCC 51299 | FA1  FA2 | ACTTATGTGACTAACTTAACC  TAATGGTGAATCTTGGTTTGG | 360 | (Molechan et al., 2019) |

**Table S2:** Antibiotic susceptibility profiles of *E. faecalis*

^a^. Antibiotic susceptibility tests were interpreted according to CLSI resistant breakpoints (v 7.1) for *E. faecalis*.

| Isolate | \| \| Antibiotic susceptibility testing ^a^ | | | | | | | | | | | | | |
| --- | --- | --- | --- | --- | --- | --- | --- | --- | --- | --- | --- | --- | --- | --- |
| Strain ID | VAN | TEC | CIP | LEV | GEN | STP | TET | ERY | CHLO | RIF | F300 | LZD | PEN | AMP |
| 1MPA1 | I | I | I | S | S | S | R | I | S | I | S | S | S | S |
| 1MPA3 | I | S | I | S | S | S | R | I | S | I | S | S | S | S |
| 1MPD4 | I | S | I | S | S | R | R | R | R | R | S | R | S | S |
| 1MPF1 | I | S | R | S | S | S | S | I | S | R | S | I | S | S |
| 1MPF3 | I | S | R | S | S | S | R | I | S | I | S | R | S | S |
| 1MPJ101 | I | S | S | S | S | S | R | I | S | I | S | R | S | S |
| 1MPK2 | I | I | I | S | S | S | R | I | S | I | S | S | S | S |
| 1MPK3 | I | S | S | S | S | S | I | I | S | I | S | S | S | S |
| 1MPK4 | I | S | I | S | S | S | R | I | S | I | S | S | S | S |
| 2MPJ104 | I | S | I | S | S | S | R | I | S | I | S | I | S | S |
| 3MPH1 | I | S | R | S | S | S | R | R | S | I | S | S | S | S |
| 3MPJ101 | I | S | I | S | S | S | S | I | I | R | S | R | S | S |
| 2UIJ104 | I | S | R | S | S | S | R | I | S | R | S | R | S | S |
| 2UIK2 | I | S | I | S | S | S | R | I | S | R | S | R | S | R |
| 2UIK3 | I | S | I | I | S | S | R | R | S | I | S | S | S | S |
| 2UPA3 | I | S | R | I | S | S | I | R | R | S | S | I | S | S |
| 2UPC4 | I | S | I | I | S | S | I | I | S | S | S | S | S | S |
| 2UPF4 | I | S | I | S | S | S | I | I | I | R | I | R | S | S |
| 2UPJ202 | I | S | I | S | S | S | S | I | S | I | S | S | S | S |
| 3UIA2 | I | S | I | S | S | S | R | I | R | R | S | S | S | S |
| 3UIC1 | I | S | I | S | R | R | R | R | R | R | S | S | S | S |
| 3UIE2 | I | S | I | S | S | S | R | I | I | R | S | R | S | S |
| 3UIJ202 | I | S | I | S | R | R | S | R | R | R | S | R | S | S |
| 3UPF3 | I | S | S | S | S | S | R | R | S | R | S | R | S | S |
| 3UPF4 | I | S | R | I | S | S | R | R | S | R | S | R | R | S |
| 3UPH1 | I | S | I | I | S | S | R | I | I | R | S | R | S | S |
| 3UIC2 | I | S | I | I | R | R | R | R | R | R | S | I | S | S |
| 1CIB1 | I | S | I | R | S | S | R | R | I | R | S | S | S | S |
| 1CID1 | I | S | S | S | S | S | R | R | S | R | S | R | S | S |
| 1CIH3 | I | S | R | S | S | S | R | R | S | R | S | S | S | S |
| 1CPK2 | I | S | I | S | S | S | R | R | I | I | S | S | S | S |
| 1CPK3 | I | S | S | S | S | S | R | R | I | I | S | S | S | S |
| 2CPF3 | I | S | I | S | S | S | R | I | I | I | S | R | S | S |
| 2CPH2 | I | S | I | S | S | S | R | R | I | R | S | R | S | S |
| 3CPH1 | I | S | I | S | S | S | R | I | I | R | S | S | S | S |
| 2SPJ101 | I | S | I | S | S | S | R | R | R | I | S | R | S | S |
| 2SPL2 | I | S | I | S | S | S | R | R | R | I | S | R | S | S |
| 2SIL2 | I | S | S | S | R | R | R | R | R | R | S | S | S | S |

**Glycopeptides**: VAN = Vancomycin, TEC = Teicoplanin;

**Quinolones**: CIP =ciprofloxacin, LEV = levofloxacin;

**Aminoglycosides**: STP = streptomycin, GEN = gentamycin;

**Tetracyclines**: TET = tetracycline;

**Macrolides**: ERY = erythromycin;

**Amphenicols**: CHLO = chloramphenicol;

**Rifamycin**: RIF = rifampicin;

**Nitrofurans**: F300 = nitrofurantoin;

**Oxazolidinones**: LZD = linezolid;

**Penicillin**: PEN = penicillin G; AMP = ampicillin.

**R, S and I =** Resistant, Susceptible and Intermediate respectively.

**Table S3:** Distribution of 6 major insertion sequence (IS)/transposase families and their associated predicted sources among *E. faecalis* isolates via the ISFinder database.

| IS Family | Number of occurrences | Predicted Sources |
| --- | --- | --- |
| IS3 | *21* | *Enterococcus faecium*  *Streptococcus agalactiae* |
| IS5 | *16* | *Cyanotheca sp.* |
| IS1595 | *15* | *Bacillus subtilis* |
| ISL3 | *9* | *Streptococcus mutans*  *Streptococcus thermophilus* |
| IS607 | *9* | *Campylobacter sp.*  *Virus NY2A* |
| Tn3 | *7* | *Bacillus thuringiensis* |
